# What has changed in the experiences of people with mental health problems during the COVID-19 pandemic? Findings from follow-up interviews using a coproduced, participatory qualitative approach

**DOI:** 10.1101/2021.08.12.21261913

**Authors:** Prisha Shah, Jackie Hardy, Mary Birken, Una Foye, Rachel Rowan Olive, Patrick Nyikavaranda, Ceri Dare, Theodora Stefanidou, Merle Schlief, Eiluned Pearce, Natasha Lyons, Karen Machin, Tamar Jeynes, Beverley Chipp, Anjie Chhapia, Nick Barber, Steven Gillard, Alexandra Pitman, Alan Simpson, Sonia Johnson, Brynmor Lloyd-Evans, On behalf of the NIHR Mental Health Policy Research Unit Covid coproduction research group

## Abstract

**Purpose:** We sought to understand how the experiences of people in the UK with pre-existing mental health conditions had developed during the course of the COVID-19 pandemic.

**Methods:** In September-October 2020 we interviewed adults with mental health conditions pre-dating the pandemic, whom we had previously interviewed three months earlier. Participants had been recruited through online advertising and voluntary sector community organisations. Interviews were conducted by telephone or video-conference by researchers with lived experience of mental health difficulties, and explored changes over time in people’s experience of the pandemic.

**Results:** We interviewed 44 people, achieving diversity of demographic characteristics and a range of mental health conditions and service use among our sample. Three overarching themes were derived from interviews. The first theme “Spectrum of adaptation”: to difficulties in access to, or the quality of, statutory mental health services, through developing new personal coping strategies or identifying alternative sources of support. The second theme is “Accumulating pressures”: from pandemic-related anxieties and sustained disruption to social contact and support, and to mental health treatment. The third theme “Feeling overlooked”: A sense of people with pre-existing mental health conditions being overlooked during the pandemic by policy-makers at all levels. The latter was compounded for people from ethnic minority communities or with physical health problems.

**Conclusion:** Our study highlights the need to support marginalised groups who are at risk of increased inequalities, and to maintain crucial mental and physical healthcare and social care for people with existing mental health conditions, notwithstanding challenges of the pandemic.

## Introduction

Since the World Health Organization declared COVID-19 a pandemic in 2020, the consequences for human health and society continue to be experienced globally. The social detriments of the virus and the restrictions put in place to reduce its spread include increased poverty, unemployment, and domestic violence, which all present continued stressors for mental health [1]. However, little has been published about how the ongoing impact of the pandemic is experienced by people with mental health conditions.

Research conducted following initial “lockdown” measures, which prohibited all but essential activities outside of the home, reported international increases in population levels of anxiety, depression and other common forms of mental distress [2–7]. People with pre-existing mental health conditions were identified as particularly vulnerable to negative psychological outcomes [2,6,8,9], as were women and young people [3,5–9]. Two large UK national surveys [10,11] reported conflicting findings about whether anxiety and depression increased for people with existing mental health conditions during the early months of national lockdown. These surveys suggest that the impact of the pandemic on people with mental health conditions may not be uniform and is not yet fully understood: qualitative exploration of people’s experiences is needed. Existing qualitative studies suggest that for some people, previous experiences of coping with adversity have been protective from the negative psychological effects of the pandemic; while for others the pandemic has worsened pre-existing difficulties [12–14]. For example, a study of people with eating disorders in the UK found that the pandemic had been a trigger for either recovery or problematic eating patterns [14]. Negative impacts for people with existing mental health conditions have included reduced access to mental health services, social isolation, and disruptions to daily routine or normal coping strategies. Reported benefits include reduced social pressures and increased engagement with recovery-promoting activities [12,13]. Unequal impacts of the pandemic were also identified among people with existing mental health conditions [12], with additional hardships amongst Black, Asian, and minority ethnic (BAME) communities, those with physical health conditions and socioeconomic disadvantages [12]. These hardships are likely to persist with ongoing restrictions. Changes to mental health service delivery, including transition to remote technologies [15], have remained in place and may continue to influence the experiences of people with mental health conditions.

Most existing qualitative studies have investigated the experiences of people with mental health conditions at early stages of the coronavirus outbreak. However, this fails to capture the impact of prolonged experiences of the pandemic, ongoing restrictions on day-to-day living, and reduced or changed access to mental healthcare and social support [12, 15], necessitating a longitudinal approach to capture prolonged experiences. To date, no qualitative research has explored the experiences of people with mental health conditions over the course of the pandemic. This paper addresses this gap by reporting findings from second interviews with participants from our previously published participatory, qualitative interview study which was based on interviews conducted between May and July 2020 [12]. The follow-up interviews reported in this paper took place three months later, following changes in many areas of England to permit social contacts and the opening of most non-essential shops, and the introduction of compulsory face coverings, but with social distancing restrictions still in place. However, localised lockdowns also took place during this time. Our aim was to explore whether participants’ day-to-day experiences and mental health difficulties had changed or stayed the same for participants since their first interview.

## Methods

We took a coproduced, participatory approach to conducting qualitative interview research, as developed and described in full in our previously published paper, with a research team including people with experience of using or working in mental health services [12]. In our initial recruitment, we recruited people with pre-existing mental health problems using targeted recruitment materials through community organisations, mental health networks, and social media. We used purposive sampling to reflect diversity of experience based on participants’ diagnosis, use of mental health services, and demographic factors, and to reflect a range of rural and urban areas. Participants were recruited to that study between 7th May and 8th July 2020, and as part of the informed consent process they were asked if they would like to take part in a second interview. All participants in the original study who stated they’d like to take part in a second interview, were approached for the follow-up interview by a member of the research team and invited to re-affirm audio-recorded verbal informed consent prior to being interviewed a second time. Interviews were undertaken by nine members of the research team working from a perspective of lived experience of mental distress and of using mental health services, namely Lived Experience Researchers (LER), supported by researchers who recorded and saved the interview securely directly onto the UCL cloud-based secure storage system. Where possible, the follow-up interview was conducted by the researcher who conducted their original interview, using videoconferencing or freephone options within Microsoft Teams. The interview topic guide sought to explore changes in participants’ experiences of the COVID-19 pandemic in relation to their mental health, ongoing experiences, and new experiences identified by participants since their first interview. The topic guide is provided in the supplementary materials.

Interview recordings were transcribed verbatim by researchers within the Division of Psychiatry or an external transcription company and anonymised prior to coding. Thematic analysis followed a modified version of the process described in detail in the original study [12]. We used the final thematic framework reported in our first study [12] as a starting point for analysis. Where possible, preliminary coding of eight interviews was undertaken by the LERs who had undertaken each first interview, as they were well-placed to identify points of continuity and changes in each interviewee’s experience. LERs coded data that articulated change, or a lack of change, in the range of experiences participants had identified in the first study, as well as coding aspects of participants’ new accounts not captured in the original framework. A revised thematic framework was then coproduced discursively with the wider research team. LERs used the revised framework to code the full set of interviews, proposing additional codes where data did not fit the framework.

Through this process, eight descriptive or semantic-level themes were developed. Summary reports of these preliminary themes are provided as supplementary material. In subsequent wider team meetings, and through a process of reviewing these descriptive theme summaries, three interpretative themes were developed collaboratively, that provide a more latent analysis. Figure 1 shows how the eight descriptive themes mapped onto the three interpretative themes. These are reported below.

**Figure 1:**
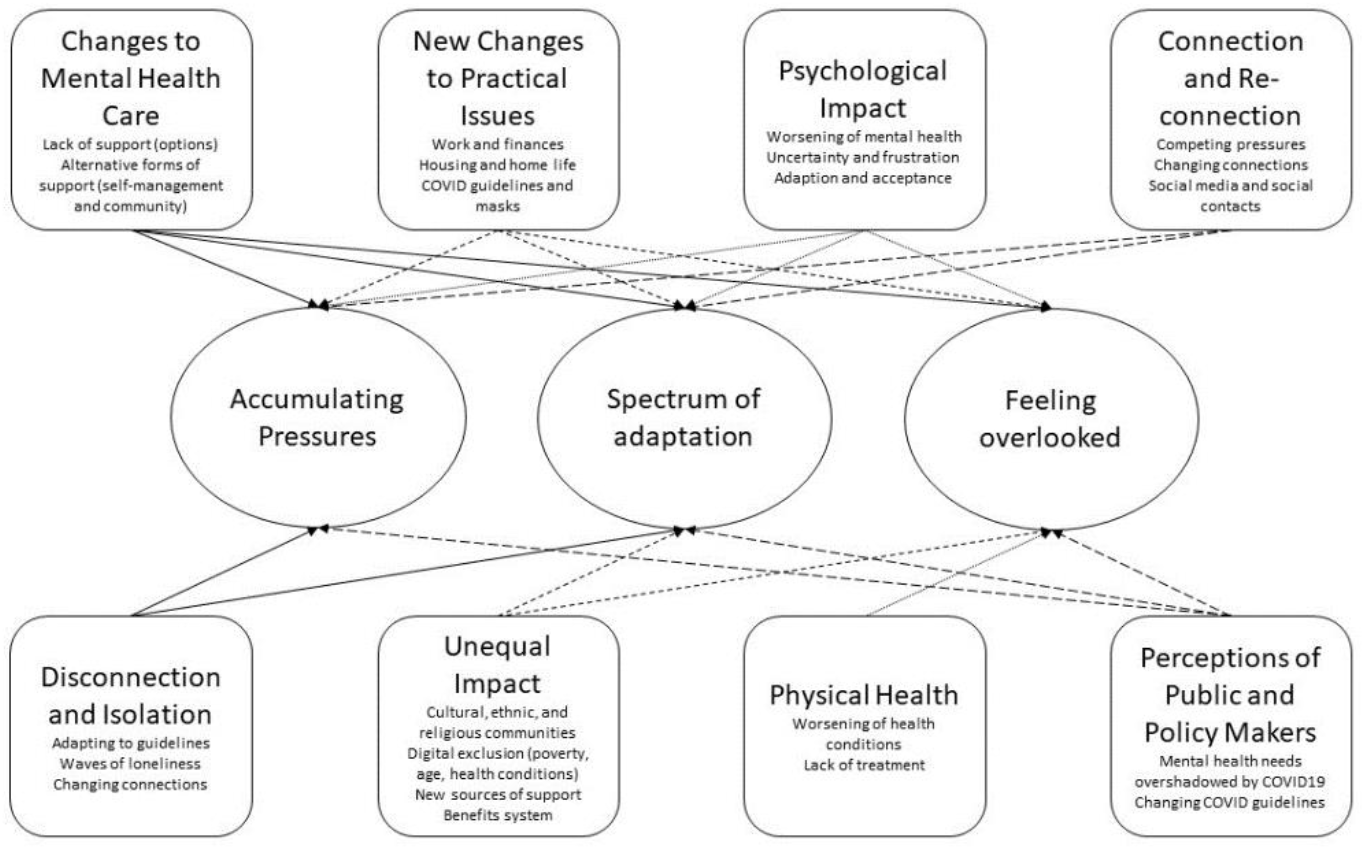
Descriptive themes mapped onto three interpretative themes reported

Ethical approval for a study focusing on loneliness and mental health problems was originally obtained from the UCL Research Ethics Committee on 19/12/2019 (ref: 15249/001). An amended topic guide covering experiences of COVID-19 and follow-up interviews was approved on 04/05/2020, and an amended topic guide for the follow-up interviews reported here was approved on 14/08/2020.

## Results

Forty-four of our original sample of 49 participants took part in a second interview. Three people could not be contacted and two declined. Follow up interviews were conducted between September and October 2020. The characteristics of follow-up interview participants are reported in Table 1.

**Table 1:** Demographics of participants (n=44)

| Characteristic | Category | Number (%) |
| --- | --- | --- |
| Gender | Female | 32 (73%) |
|  | Male | 12 (27%) |
| Age (years) | 18-25 | 3 (7%) |
|  | 26-35 | 14 (32%) |
|  | 36-45 | 13 (29%) |
|  | 46-55 | 6 (14%) |
|  | 56-65 | 3 (7%) |
|  | 66-75 | 3 (7%) |
|  | Information not available | 2 (4%) |
| Ethnicity | White British | 24 (54%) |
|  | White Other | 4 (9%) |
|  | Mixed/Multiple Ethnic Groups | 3 (7%) |
|  | Asian/Asian British | 6 (14%) |
|  | Black/Black British | 6 (14%) |
|  | Other Ethnic Group | 1 (2%) |
| Sexuality | LGBTQI<sup>1</sup> | 8 (18%) |
|  | Heterosexual | 30 (68%) |
|  | Prefer not to answer or information not available | 6 (14%) |
| Region of UK | North (North East, North West, Yorkshire and Humber) | 8 (18%) |
|  | Midlands (West Midlands, East Midlands) | 5 (11.5%) |
|  | South (South East, South West and East of England) | 5 (11.5%) |
|  | London | 25 (57%) |
|  | Wales | 1 (2%) |
| Urban/Rural Location | City | 35 (80%) |
|  | Town | 8 (18%) |
|  | Village | 1 (2%) |
| Employment | Full-time paid employment | 10 (23%) |
|  | Part-time paid employment | 10 (23%) |
|  | Furloughed<sup>2</sup> | 3 (7%) |
|  | Not in paid employment | 21 (47%) |
| Current mental health Service Use | None or on waiting list | 13 (29.5%) |
|  | NHS Community mental health services<sup>3</sup> | 22 (50%) |
|  | Crisis house | 1 (2.3%) |
|  | GP or Primary Care counselling | 5 (11.4%) |
|  | Private sector psychotherapy only | 1 (2.3%) |
|  | Voluntary sector mental health services only. | 5 (11.4%) |
| Self-Reported diagnosis | Personality Disorder | 6 (14%) |
|  | Mood Disorder (Depression, Anxiety, PTSD) | 19 (43%) |
|  | Bi-polar Disorder | 6 (14%) |
|  | Schizophrenia/Psychotic illness | 5 (11%) |
|  | OCD | 1 (2%) |
|  | Not stated | 7 (16%) |

<sup>1</sup> LGBTQI included: Gay (n= 1), Lesbian (n= 1) Bisexual (n= 5) Pansexual (n=1) 2. Furloughed: temporarily laid off from work due to the pandemic, with wages still paid in part by a government assistance scheme 3. Community Mental health services included: Community Mental Health Team (n=16) Therapist (n=5) NHS Peer Support Service (n=1)

Three overarching, interpretive themes are presented below with sub-headings representing concepts within those themes rather than explicit sub-themes.

### Theme one: Spectrum of adaptation

Reflecting on the period since the first interviews three months earlier, participants reported a wide spectrum of experiences in learning or being forced to adapt just to cope with the ongoing pandemic. However, not everyone reported being able to adapt successfully.

#### Finding alternative forms of support

While many participants described reduced access to or quality of support from NHS mental health services during the pandemic, many participants had looked for alternatives to support their mental health, including online support groups, digital forums and social media, apps, meditation, art therapy and other activities. Use of the voluntary sector, community, and faith-based support, as well as self-management, became increasingly important:

> *“I use Mind because I’m comfortable with them, because I do volunteering with them and I’m very familiar with the people… so I’d rather use their services than any other service*.*’’* [P10]

However, although use of these alternative forms of support was helpful for many people, for some there was a sense of having to rely on them because no other options were available – a forced adaptation – rather than having benefited from connecting to a wider array of support sources. Several participants who felt let down by statutory services had been unable to find or develop suitable alternatives to care, or new ways of coping in the absence of formal help.

> ‘‘*I attempted to download a mental health app, but it was free and wanting my information, so I didn’t trust it, so I then uninstalled it*.*’’* [P27]
>
> ‘‘*I tried to do [a group] with Mind but I just found it really negative. I feel maybe the people in the group were maybe in quite a significantly worse state possibly or even the way the facilitator was structuring it, it just all felt a bit doom and gloom. I think I did one and then I left it be*.*”* [P41]

#### Personal adaptation strategies

In addition to finding external forms of support, some people developed new routines, creative and active pursuits, and self-care strategies to cope with daily life, adjusting these as the pandemic continued. Participants reported adapting their home environment as well as acquiring new pets, which had a positive impact on mental health.

> *“[The new puppy is] something else to focus on and forces me out of the house. So, it definitely was a conscious decision in terms of improving my mental health and giving me purpose and structure, which I didn’t have before*.*”* [P32]

#### Adaptations in human contact and digital connections

Participants frequently observed that lockdown had initially led to a welcome relief from personal and social pressures to see others, particularly if previous social interactions had tended to raise individual anxieties. However, for some, their loneliness had increased with continued absence of human contact leading to further feelings of isolation, highlighting an often unmet need to interact with other people.

Digital connections had been helpful during the lockdown, and people continued their use of video call platforms, including to speak with friends and family globally.

> ‘‘*A lot of my friends are like scattered around the country, or where I grew up or where I studied before, so it’s kind of, in a cheesy way, not closer to them but it’s been nice to mix the friendship groups a bit more and get the home girls together on a Skype, which we would normally never do*” [P3]

Some people had become more boundaried and mindful about their use of social media, having learned their limits in balancing the downsides *versus* benefits and subsequent mental health impacts.

> “*I have dropped out of social media, so I am having less of those surface friendships. More trying to focus on the friends that I can see, and that we spend time together rather than just commenting on each other’s Facebook statuses*.*”* [P19]

Participants also described adapting the degree of in-person contact in response to the changing guidelines, such as forming support bubbles and being able to hug, and their positive impacts:

> “*when we were allowed to form the support bubbles, that was really good for me. I was able to start going and having lunch with my mum. My mum and I formed our support bubble and we started doing Sunday lunch again. Every other weekend we were doing a movie night. Just things like that, which was really lovely, just to be around somebody again and actually hug someone*…*”* [P23]

### Theme two: Accumulating pressures

Participants described feeling mounting pressures caused by sustained exposure to the pandemic restrictions, trying to manage conflicting needs, and changing personal circumstances, as well as the inconsistencies or sudden changes they perceived in local and national restrictions. This was commonly experienced as a subtle ratcheting of pressure:

> *“It certainly feels as if there has been a tightening of the noose”* [P11]

#### Changing circumstances

Factors such as changes during the pandemic to the support provided by mental health services, people’s social relationships, financial circumstances, and lockdown restrictions and government guidance all served to increase pressures on some participants. One participant described frustration, anticipation, then dismay at trying to re-establish support from mental health services.

> *‘‘I obviously got seen, and reviewed, and then discharged in one appointment’’* [P21]

Another participant described no change in contact with mental health services or GP as he had been discharged before the first lockdown and had not been able to see his GP to ask them to follow up with mental health services.

> “*I am in exactly the same situation [having been discharged by the community mental health team just before the first lockdown]. There has been no development with them. I actually don’t even know… I haven’t seen my GP to be able to follow that up*…*I saw him just once shortly after the lockdown to do some blood tests, but… I think I saw him a couple of times, but that was at the very beginning of lockdown and I have not seen him since*.” [P40]

Another participant reported that their partner was struggling with lockdown and there was a role reversal in supporting them after they had supported her for years: *“it’s my turn now”* [P48]. Others described anxieties resulting from increasing financial pressures and a new need to engage with the benefits system:

> *‘‘And it has sort of got to the point where I can’t pay the rent in its entirety”* [P13]

Since the previous set of interviews, a national easing of restrictions had enabled many participants to meet up with others in person, and this was felt to have had a positive impact on mental health. However, many participants referred to the emotional impact of changes to the levels of permitted contact with family and friends. One interviewee described how it felt to have restrictions lifted but then reimposed locally:

> *“It was good while it lasted. It was very positive to be able to see a few people during the time where we could. I think it’s more frustrating having had it taken away than perhaps if we’d never had it in the first place*.*”* [P22]

Trust in the government appeared to have eroded since the first set of interviews: many described official guidance as illogical, inconsistent and badly communicated. This was perceived as anxiety-provoking and left many feeling isolated.

> *“They* [the government] *increase anxiety, they increase my anxiety, they*…*just keep chopping and changing with things, which means that you can’t tell what’s good or what’s not. All of their advice seems to be conflicting*’’ [P27]

Despite some participants starting to venture out again by using public transport following the introduction of legislation to wear masks on public transport, which supported their mental health, other participants reported increasing negative mental health impacts of compulsory mask wearing:

> *“I find that wearing a face mask has a negative impact on my anxiety levels*…*so we are all wearing face masks, and my anxiety is actually very bad at the moment. In college it’s really bad. Everytime I go to talk, I have a panic attack*.*”* [P8]

#### Conflicting needs

Over time, responding to these pressures created conflicting needs, for example for activity and social contact *versus* minimising infection risks.

> *‘‘I was very near self-harm. I cannot sit in my flat all day on my own. Even if I’m going to die of COVID, I have to go out for a bit*.*”* [P6]

Whichever course of action participants decided on was seen to potentially threaten mental wellbeing. *“I did eat out a few times, although, again, it was with caution, and I was scared…. I thought, “God, I might just die because of this half-price thing*.*”* [P42]

> *“Sometimes it’s better to be in your bedroom but it’s not healthy and I’m losing skills, I’m losing friends”* [P48]

Some participants also expressed feeling conflicted about whether to access much-wanted mental health support. One felt that she was not a priority for help:

> *‘‘Part of me feels I don’t desperately need it, and also I know when things are stretched and at these stressful times there are definitely gonna be so much more desperate people that need it way more’’* [P4]

#### Cumulative impact

Participants’ experiences of changes in their mental health during the pandemic were not uniform. A few reported improvements in wellbeing:

> *“Luckily, I went into lockdown in like a good place, even though I’ve been up and down on lockdown, I don’t think it’s been as bad as pre-lockdown’’* [P3]

However, many people reported a progressive deterioration in their mental wellbeing, attributed to factors including: lack of contact with family, friends and society; worry about loved ones’ health; the absence of support from mental health services; frustration over guidelines and others’ non-adherence to them. and loss of optimism and hope of life ever returning to normal. Reported mental health difficulties included a worsening of depressive symptoms, paranoia and anxiety:

> *‘‘I’m just getting so depressed. But my mind is getting distorted with the horror of this feeling. This feeling that I’m completely abandoned and it’s going to go on forever. I do feel like I don’t want to live like that*.*’’* [P6]
>
> *“My symptoms have got a bit worse. My paranoia has got worse. I can hear people talking about me, and I’m sure it’s not real*.*”* [P8]

People described feelings of being in limbo, a sense of uncertainty, and frustration from ongoing exposure to the pandemic and restrictions, where accumulating pressures increased mental vulnerability, and made it hard to maintain hope of life returning to normal.

> *‘‘ If something negative happens, it’s probably hitting me harder than it used to*.*’’* [P9]
>
> *“But I’m not sure that the outbreak is temporary. I think that may be semi-permanent. That’s what gets me quite scared”* [P11]

### Theme Three: Feeling overlooked

Participants reflected that, as described in the first interview, the government’s response to the pandemic lacked consideration of the specific needs of people with mental health problems. For people from marginalised communities in particular, this compounded a sense of being overlooked by government and societal responses to the pandemic compared to the general population.

#### People with mental health problems were not a focus

Several participants felt that the government guidance and restrictions did not take account of those with pre-existing mental health issues. They expressed the view that this blinkered policy-making conveyed a lack of understanding of the needs of people with mental health problems that heightened the detrimental impact of isolation on these groups.

> *“I think they [policy-makers] get side-lined by Covid to the point where they forget that people have mental health problems*.*”* [P46]

Participants felt frustrated by apparently mixed messages, for example that it was safe to travel to work or socialise, but not to see a mental health professional with infection controls in place.

> “*How comes someone can go to the pub, but I can’t see my therapist?*” [P4]
>
> “*It’s the one thing that has changed in my opinion of the government since your last interview, because I’ve got angrier at them. So, I am just going with my own instincts, going with what the experts are saying*” [P27]

Participants voiced frustrations about navigating benefits systems for the first time during the pandemic and felt the needs of those with mental health problems were not being recognised, particularly given the benefits system being perceived as unduly complicated.

> *‘‘I have been looking into benefits, which has been annoyingly complicated. Universal Credit is a mess, as far as I can tell, and trying to look at any benefits I might be eligible* …*that hasn’t been good, at all… there is a gap there, as far as I can see, for people who suffer with mental health problems*.*’’* [P26]

#### Health services

For some participants, their access to mental health services had improved since their first interview. However, others felt mental health services were erratic, hard to navigate, or not responsive to needs. Lack of investment in mental health services was mentioned as something that heightened these perceptions. Participants conveyed a perception of a lack of duty of care within the mental health system, as well as services not being person-centred, there being a lack of options, and a sense of having to fit into the service rather than vice versa.

“*I need to be speaking to somebody, like a coordinator, who’s going to show a bit of compassion, once every two weeks, and I’m not getting that. Really, I need to see the psychiatrist, and not for it to be open-ended, when the next appointment is, or if I’m even getting another appointment. It needs to be structured*.” [P9]

One participant described how appointments were increasingly difficult to arrange:

> “*In terms of official mental health services, they have just not been very responsive or available during the pandemic. If I have an appointment with the doctor or the psychiatrist, then I know never to cancel those. Because they are like gold-dust”* [P35]

For some participants, the groups run by mental health services that they attended to support their mental health pre-March 2020, had not resumed at the time of the second interviews, and others had ongoing difficulty accessing specific services:

> “*I have also had a lot of problems with the personality disorder unit. I haven’t seen them face to face since March, and I have raised it lots of times, over and over and over. Although in part that is because my care coordinator is working from home, but in part it is just because of whatever their own agenda is*.” [P44]

There were also difficulties reported in accessing physical healthcare because of anxiety regarding using public transport, and of Covid overshadowing needs for appropriate physical health checks. This was particularly anxiety-provoking given well-publicised information about the extra threat of Covid to those who were overweight, had heart disease or diabetes, and the impact of lockdown on general health, for example, reduced exercise and weight gain experienced by some participants.

#### Cultural needs unheeded

Some participants expressed perceptions of shrinking emotional and practical support systems within their community. Participants thought the government overlooked the religious needs of groups such as Muslims, particularly in its handling of restrictions over Eid. This was felt to impact on the mental health of those communities specifically, given the value placed by the Muslim community on the support inherent to gathering together at Eid.

> “*For example, Eid, because of being Muslim. We have had two Eids now, where we couldn’t even celebrate them together. Waiting the night before the second Eid, to find out whether we would be allowed or not*… *I think, on the government’s part, that was really unfair. If it had been Christmas Eve, I don’t think they would have left it so late, put it that way*.*”* [P23]

#### Economic and digital needs overlooked

The government’s reliance on digital access to food, education, and social connections was felt to ignore those digitally excluded through poverty or age group due to a lack of access to equipment or the skills to use it. Older populations felt particularly left behind by the quick transition to online services.

> “*And then you’ve got internet poverty and not having the right equipment or having the right speed or having the right tech and stuff like that*.” [P17]

## Discussion

### Main findings in context

Our findings highlight the particular and ongoing struggles of those with pre-existing mental health problems during the COVID pandemic, and the degree to which they felt forced to adapt to the lack of services available to address their mental health needs. These findings highlight the challenges for healthcare services and service users in maintaining access to physical and mental healthcare, and people’s experience of reduced access to social support and economic opportunities. The accumulation of pressures which many participants described represents a very clear threat to mental health, which could worsen existing mental health problems. The sense of injustice and abandonment conveyed by participants in feeling their needs were overlooked by policymakers at all levels was an added burden.

To our knowledge no other longitudinal qualitative studies have probed the impact of the pandemic and its accompanying restrictions on people with pre-existing mental health conditions. However, other UK qualitative studies have identified similar themes of disruption to mental health services and the personal difficulties created for people with mental health conditions by ongoing uncertainty and a sense of lack of control [13].

Our findings can also be triangulated with the quantitative findings of a longitudinal survey of the UK population, which found less favourable depressive symptom trajectories in people with pre-existing mental and physical health conditions [16].

Participants’ experiences in our study were varied. While they were negative for many, some people developed personal adaptation strategies and located new sources of support. Some people reported benefits during lockdown from reduced social contact and pressures. This mixed picture has also been reported in other qualitative studies during the pandemic: of older adults, where some participants mourned the loss of ‘normal life’ and the activities that normally protected their wellbeing, whilst others reported that the slower pace of life was protective of mental wellbeing [17]; and of people with mental health problems [13]. In our study, these positive experiences of the pandemic and lockdown were only reported by a small minority of participants. For most people, the adjustments were experienced as a forced adaptation, due to having so few other options available.

### Strengths and limitations

By following up participants from our earlier interview study [12] we addressed a gap in the literature providing an in-depth longitudinal exploration of the impact of the pandemic beyond the first UK lockdown on people with pre-existing mental health conditions. Our work provided critical detail and depth to complement the findings of longitudinal national surveys, gaining a rich understanding of the specific issues affecting people with mental health problems. The purposive sampling we used for our original interview sample achieved a diverse participant sample, and the uptake of follow-up interviews was high (44/49; 90%). The experiences of people in a range of ethnic groups were thus well represented. This is important due to the unequal mental health impacts of the pandemic on people from minority ethnic groups [18]. We also over-sampled people identifying as from a sexual minority: 18% compared to the national proportion of 4% [19]. Our sample was drawn from a wide geographical area, but will have under-represented the digitally excluded, and over-represented people living in urban areas (and London in particular). We included people with a broad range of mental health conditions and who were supported in various healthcare settings. One further limitation of this paper is that the breadth of issues covered prevented an in-depth exploration in this paper of specific issues encountered by ethnic and sexual minority groups, or groups defined by specific mental health problems.

Our coproduced, participatory approach to collecting and coding data meant that our analysis, much of which took place through regular team discussions, reflected a range of perspectives and experiences. Challenges to coding decisions improved reflexivity and improved the validity of our coding framework. Our transparent approach to coding presented our initial framework of eight themes, and how we derived three over-arching themes from this starting point. Our coding and development of themes sought to describe changes over time and developments in people’s experience of and reaction to the pandemic since the first set of interviews. We recognise however that some participants perceived little change during the period covered by these two studies, and this continuity has been less fully described in our results.

Our interviews were only able to provide a perspective on experiences of the pandemic up to October 2020: people’s experiences and responses are likely to have changed further since then, particularly with the reinstitution of national lockdowns in England in November and December 2020. The geographical diversity of our sample complicates the process of locating these experiences in context, as participants were experiencing different degrees of local lockdown at the time of interview.

### Clinical and policy implications

Our research study presents policy-makers and commissioners with a valuable needs assessment for this vulnerable group, suggesting that it is unsustainable to expect people to rely almost solely on digital resources (online support groups, digital forums, social media, apps), voluntary sector support, religious communities, or self-care to manage mental health conditions which predate the pandemic. Whilst some had found these new sources of support acceptable, more had found a forced adaptation to these resources inadequate. Even in our study sample, which will not have included the most digitally excluded, tele-mental health care was problematic for some, due to lack of necessary equipment and connectivity, or lack of appropriate privacy in their home environment.

Our work shows that policy-makers and service planners need to prioritise maintaining and optimising access to mental and physical health care for people with existing mental health conditions, as this was a concern expressed by our interviewees. This will require initiatives to address digital exclusion of those with mental health problems and optimise the implementation of tele-mental health care, as advocated in national policy guidelines [20, 21]. It may also require a more proactive approach to maintaining safe face-to-face assessment and treatment of mental and physical health during the pandemic and beyond, as well as better initiatives to address social connectedness. Voluntary sector organisations and local community groups may be well-placed to provide agile, bespoke responses to developing local needs [22] but are also often constrained by social distancing and online capability and may find it hard to access or retain funding during periods of economic uncertainty [23]. The impact of COVID and pandemic restrictions on people with mental health problems might be regarded as a civil rights issue [24], requiring governments to focus their resources on redressing the inequalities and compounding intersectional inequalities we observed.

### Further research

With virus mutations, and despite the roll-out of the vaccination programme internationally, further waves of infection and cycles of restrictions are possible. These, together with its long-term economic impacts, suggest the pandemic is likely to have sustained and potentially increasing mental health impacts [1]. Therefore, further qualitative research, including longitudinal follow up studies, is needed to understand people’s developing experiences over time and the challenges they are facing. This should include a focus on specific groups among those with mental health conditions whom we identified as feeling overlooked and facing challenges, including the digitally excluded, people with comorbid physical health conditions, and people from minority ethnic communities. Understanding people’s coping responses to the pandemic and how these may help or hinder wellbeing over time is also of high importance: ongoing cohort studies can help identify predictors of worsening mental health problems during the pandemic [25]. Further quantitative work using anonymised health records can help identify key correlates of access and inequalities in access to mental health care. Emerging findings [26] indicate a widespread shift to remote consultations and a reduction in access to community mental health crisis care during the early months of the pandemic., and this is likely to have an unequal impact, worsening health inequalities.

### Conclusions

As the COVID restrictions continued throughout 2020, our longitudinal follow-up of a UK sample of people with mental health problems describe a ‘tightening noose’ of pressures and the restrictions on access to formal and informal support sources, and the impact of these pressures on people’s pre-existing mental health difficulties. Despite being forced to find ways to adapt to COVID restrictions, both during lockdown and in the period after this, many participants in our study reported struggling to cope and a deterioration in psychiatric symptoms. Policy responses should seek to optimise tele-mental health, to offer personalised options for care delivery that meet people’s needs and preferences, and strategies to reduce digital exclusion, including alternatives to remote healthcare where required. Our study highlights the need to reach and adequately support further marginalised groups who are at risk of increased inequalities and maintain crucial mental and physical healthcare and social care for people with existing mental health conditions, notwithstanding the logistic and financial challenges of the pandemic.

## Supporting information

Descriptive Themes analysed to develop final three themes

Follow-up Interview Topic Guide

## Data Availability

Interview transcripts are not publicly available to ensure individual participants are not identifiable in accordance with participants' consent and study ethical approvals.

