## Supplementary material for "What has changed in the experiences of people with mental health problems during the COVID-19 pandemic? Findings from follow-up interviews using a coproduced, participatory qualitative approach": Descriptive Themes analysed to develop final three themes

**Title of Study:** Exploring the lived experiences of loneliness and isolation with people with mental health problems during the COVID 19 pandemic in the UK

**Eight Descriptive Themes analysed to develop final three themes**

**Changes to mental health care: Theme summary 1**

Some participants were still not getting access to mental health support in the way they would like, such as via face to face and more frequent appointments. There was still limited access to types of support therapy and groups. However, some people had more positive experiences of statutory services, for example below:

*“So they said, when I was there after my A&E trip, they said in the future, if you don't need to go to A&E for some physical thing, you can come straight here for, like, a psych assessment. And so they set up a, kind of, psych A&E at that hospital, which I thought was... I thought at the time, I must remember that and use it if I need to, but I haven't actually needed to since. And I don't know if that was just for the height of COVID and whether it's still going, but I think that was quite a good idea to have somewhere that wasn't A&E that you could go to just to be assessed and talk to someone and see if you could get some help with stuff.“* [P1]

Many services were not being person-centered, as there was a lack of offering different support options, and it seemed more about the participant fitting to the service, not the service fitting to the participant. This theme continued from the first interviews, but came across more strongly this time:
*"I need to be speaking to somebody, like a coordinator, who's going to show a bit of compassion, once every two weeks, and I'm not getting that. Really, I need to see the psychiatrist, and not for it to be open-ended, when the next appointment is, or if I'm even getting another appointment. It needs to be structured.”* [P8]

Some participants described feeling let down by CMHT, or GP. Lack of continuity of care came across quite strongly, for example having to speak to locum at CMHT.
This was a strong theme across participants and many participants had looked for support elsewhere, or alternative ways for them to support their mental wellbeing. It seemed alternative support had widened up more, such as: support groups – formal and informal, forums, social media, meditation, yoga, art therapy, which were all online. A few participants had used apps, which was not really spoken about during the first interviews. One participant was enthusiastic about discovering the monthly online forums hosted by ‘We Coproduce’ and there was a sense of being part of a community.

Participants were also being proactive themselves in finding support outside of statutory services in order to support their mental wellbeing. Therefore, although things were challenging participants were showing how resilient they were. It came across strongly that voluntary sector services, such as Samaritans and Mind, were continuing to help many participants.

*"I use Mind because I’m comfortable with them, because I do volunteering with them and I’m very familiar with the people… so I’d rather use their services than any other service’’* [P10]

### **New Changes to Practical Issues: Theme Summary 2**

Although some had now adopted to working from home, others were still struggling with the idea of working from home as it made some feel isolated while others felt it took away their routine.

*‘’I’m not somebody who could work from home. I find being at home very isolating...I just don’t feel that it would be good for my mental health to work from home.”* [P25]

Finances

Some participants reported pandemics impact leading to reduced finances since the last interview (e.g., due to losing work or having benefits assessments paused), continued lack of support, threat of further lockdowns and potential job losses. This highlighted inequalities as individuals with mental health needs have been harder hit now more than the previous interviews.

“*Especially the housing situation, because that’s so precarious right now, they will cut housing benefits and we won’t be able to basically live.’’* [P46]

Those working in support roles had to manage their own mental health as well as others mental health alongside aspects of uncertainty of job security and growing demands to work differently.

*‘’The uncertainty and anxiety that everyone has been feeling about people’s jobs... and the panic of trying to get everything online...’’* [P3]

Many participants noted ways that they had adapted their home life to help cope with ongoing lockdowns. Since the last interview, several participants had adopted pets greatly improving their mental health and reducing feelings of isolation:

*‘“I've got a new puppy. I don't know if that applies, but, in terms of humans, I'm the only one and still living in the same flat... he has been such an improvement to my mental health...”* [P32]

For those individuals who previously did not have time to undertake home improvements, they suddenly found themselves this time round with time to do some DIY which also gave them a sense of accomplishment:

*‘’ We have done quite a bit of work to the house, I suppose... that has been quite nice. There has been a few jobs that have been bothering us. So, we have done some painting and stuff, you know, DIY.’’* [P26]

Conversely, others felt pressured or too overwhelmed by the pandemic stress to motivate themselves to engage such activities;

*‘’They cannot comprehend why I’m not going to paint my flat. I’m just too agitated, you know.’’* [P6]

Anxiety levels due to continued restrictions had risen for some as they continued to worry about vulnerable friends and family. One person reported that their partner was struggling with lockdown and there was a role reversal “it’s my turn now” to support them after they supported her for years. Strengthening of relationships was reported by others as a change. Not everyone reported a positive change in their homelife. For some, there was a growing sense of loneliness and living in accommodation which they felt was substandard and impacted their mental health, one participant had since weighed the risks of catching Covid-19 against that of a deterioration in their mental state:

*‘’ Because of the loneliness thing, I have discovered that it’s dangerous for me to stay inside the flat, because I do go crazy, you know. I was very near self-harm. I cannot sit in my flat all day on my own. Even if I’m going to die of COVID, I have to go out for a bit.* [P6]

The guidelines on mask wearing had caused a lot of confusion with some interviewees in previous interviews, with one stating they were not *‘’ convinced about it’’*, however, since it became a part of enforcement to wear masks in certain settings, they had since complied because *‘’on the basis of avoiding further unnecessary hassle... I am complying with the regulations.”.*

Others pointed at the practicalities of wearing masks, including steaming of glasses, sweating, and feeling breathless. For one interviewee, the biggest change around requirements of mandatory mask wearing had brought about *‘’fear of the fear’’* [P42] of catching the virus in public. In previous interview they had stopped using public transport, however, since introduction of legislation to wear masks on public transport, they had started venturing out. Other participants expressed frustration of not everyone complying with wearing masks, with one stating that they should have been medically exempt, however, still wore a mask for fear of being ostracised.

### **Psychological Impact: theme summary 3**

There were a wide range of experiences reported in changes to mental health over time during the pandemic. Many people reported that they had become increasingly depressed and anxious over time. This was attributed to the ongoing uncertainty of lockdown and the time course of pandemic; the lack of adherence to guidelines by others; the lack of in-person contact with family and friends; the enduring effects of loneliness and isolation; worry about loved ones’ health; the absence of contact and appropriate care from mental health services; and the loss of optimism and hope of life ever returning to normal. Some people reflected on how much more anxious they felt about going outside when they had stayed at home for so long.

“*I'm quite anxious about going out twice a week when I haven't been going out at all*” [P11]

Others described how their paranoia had increased:

“My symptoms have got a bit worse. My paranoia has got worse. I can hear people talking about me, and I'm sure it's not real.” [P8]

Since the first interviews, a number of people reported starting new medication for mental health, or increasing the dose. A few people described how they had reduced the dose or were managing without it entirely.

“I’ve noticed increasingly in lockdown is I’m barely taking my medication at all. I haven’t need to go and get a repeat prescription from my GP or anything or go and have a check up about it. I’ve got some medication in my bedside drawer, but I just haven’t needed to take it.’’ [P3]

Many people described a feeling of limbo, uncertainty, and frustration from the frequent changes to government guidance, and this became harder to bear as time went on, particularly the impact of isolation.

‘’I have been more worried, because I don’t really know when it’s going to end. With the new restrictions today, it’s like now we’ve become restricted again. So, how long is this going to last? One more year, two more years?” [P18]

There was also frustration at the lack of consistency in the guidelines:

“*So it’s OK for me to go into jobs in the middle of the city...but not to see my psychiatrist in a room that could be controlled* [Mask, distancing etc]” [P48]

Some people described stress and anger from seeing how other people were disregarding the infection control rules:

“*It’s caused my depression to spike quite a bit. There’s a lot of frustration around it because I know myself, my household and all of my friends take this very seriously and follow all the guidelines. The people I work with all follow the guidelines. If you go out around town you see lots of people not wearing masks in plenty of other shops before restrictions tightened.*” [P22]

However, there was also a sense of adaptation and acceptance, and for some, the new development of routines, creative and active pursuits, and self-care strategies to cope with daily life.

“*Also, it's [new puppy] something else to focus on and forces me out of the house. So, it definitely was a conscious decision in terms of improving my mental health and giving me purpose and structure, which I didn't have before*.” [P32]

### **Connection and re-connection Theme Summary 4**

The lifting of restrictions since the first interviews, allowed physical meetings with a limited number of people, and on the whole, participants welcomed this opportunity to reconnect.

Most interviewees started to go out again, including to explore new areas, return to volunteering, or start training courses. Several people mentioned going out to eat with friends and family, although one person noted the concerns this had caused:

*“I did eat out a few times, although, again, it was with caution and I was scared. Some restaurants just had the aircon on and closed all windows and I just thought, “Oh God, that’s really bad,” and I thought, “God, I might just die because of this half-price thing.”* [P42]

Digital connections had been helpful during the lockdown, and people continued their use of Zoom and other platforms, including to speak internationally with friends and family.

Some people had become more boundaried about their use of social media:

*“I have dropped out of social media, so I am having less of those surface friendships. More trying to focus on the friends that I can see, and that we spend time together rather than just commenting on each other’s Facebook statuses.”* [P19]

Many people noted the improved relationships created through increased efforts to stay connected to friends and family, and described the relief at being able to see them again.

*“Human connection is what has been lost over this virus. I think, after the measures have been lifted, people are just feeling as if they can touch each other, or they can have eye-contact or maintain distance with social-distancing measures. I think it’s just created a lot of relief on people’s minds”* [P7]

In contrast, one person suggested that when restrictions were lifted they felt more isolated because people were going back to work:

*“In normal circumstances, when everybody is at work, it’s very quiet where I live and very isolating. So, I’ve actually found the lockdown quite helpful with my mental wellbeing. …- As we gradually came out of lockdown, I started to feel more isolated again.* [P27]

While many people limited their number of contacts and complied with rules around social bubbles, there were tensions for individuals who questioned the reason for specific limits or who noticed others’ disregard for local rules, and also amongst friendship groups where some may still be shielding. Anticipation of future restrictions caused further tension:

*‘’I am torn between meeting up with friends more over the next month before things get worse, or not doing it because that would exacerbate things getting worse’’* [P27]

However, some areas of the UK were restricted again at this point, and one response describes how it felt to have had the opportunity for contact removed:

*“It was good while it lasted. It was very positive to be able to see a few people during the time where we could. I think it’s more frustrating having had it taken away than perhaps if we’d never had it in the first place.”* [P22]

### **Disconnection and Isolation: Theme Summary 5**

Since the previous interviews, a national easing of restrictions had enabled many participants to meet up with others in-person. Although a number of participants reported that this had reduced their feelings of loneliness, many were balancing fears of risk of infection with their need to connect with others to reduce the impact of isolation on their mental health:

‘*’Because of the loneliness thing, I have discovered that it’s dangerous for me to stay inside the flat, because I do go crazy, you know. I was very near self-harm. I cannot sit in my flat all day on my own. Even if I’m going to die of COVID, I have to go out for a bit.”* [P6]

For some participants, their in-person contacts continued to be limited, either physically due to local lockdowns, or through loss of contact with other vulnerable individuals who were shielding, who feared contracting the virus or were struggling with personal experience of mental distress. Many participants continued to experience isolation, in-spite of the national changes:

“*they've got progressively worse [feelings of loneliness] in that I feel more and more isolated. … hardly any human contact*.” [P11]

For some participants, these continued experiences of loneliness had a circular effect, increasing feelings of isolation since the previous interview and creating a sense of a confining state of being “*the same now, only more so*” [P11].

Similarly to the previous interview, the loss of face-to-face contacts within mental health services, workplaces or community groups often exacerbated feelings of disconnection from others:

‘’ *there are people in the workplace who they were not in my direct team. who I would bump into and have a chat with, or I would arrange to go and have lunch with. Those connections have been lost.’’* [P35]

Some participants related changes to their feelings of loneliness and isolation directly to their levels of human contact, whereas for others, loneliness was seen as a constant feature of their lives, often pre-dating the pandemic:

“*I think it [loneliness] is the same, to be honest...I think it is constant. I feel like it is something that just follows me around*.” [P29]

Participants frequently noted that lockdown had initially led to a welcome removal of personal and social pressures to see others, particularly if social interactions raised individual anxieties. However, a paradox of loneliness emerged as continued loss of human contact led to further feelings of isolation and highlighted an often un-meetable need to interact with other people:

"*You need to physically see people as well. I think that’s the thing. Certainly, for me, seeing people over the computer… ... . It just made me realise that. I always thought I could manage without people and you could send me off to a little island to go and count penguins or whatever, but actually, now, I don’t think I could."* [P30]

### **Unequal impact: theme summary 6**

Overall, the data highlighted that there were key inequalities that were impacting a range of participants within the study. Most widely cited was the continued impact that was felt by those from BAME communities were death rates within their communities continued to be higher thus created higher levels of anxiety and fear as a result of the vulnerability felt. For a few participants the shrinking support systems within their community due to lockdowns increased their feelings of isolation as they felt that due to cultural beliefs their family didn’t understand or support their mental health.

*“We don’t know how to deal with it [mental illness].”* [P10]

*“I’ve observed whenever you talk to the south Asian community, a lot of people their opinion is, “Go back to work. It’s not an illness. What are you on about? I don’t understand.”* [P7]

This was a continued issue from the first interview however the ongoing lack of services and social isolation made this feel more enhanced for some people. The importance of the support from their faith communities across all ethnicities and religions was highlighted by a few participants who noted that these became a source of positive support, both emotional and financial in one case.

*‘’ I have also been lucky enough to get to my church, and I think they said to me, “Are you alright for money?” And I said, “Well, no. Not really.” And they said, “Well, can we help you?” And I said, “Well, yes, perhaps you could.” And they have given me £150 on two occasions, which has been great.* [P13]

This positive impact was only felt where individuals could access those spaces, however restrictions in some areas meant this was limited. One participant noted that these restrictions high BAME faith communities harder as events such as Eid were impact last minute which increased their feelings of unimportance and invisibility in society.

*“for example, Eid, because of being Muslim. We have had two Eids now, where we couldn't even celebrate them together. Waiting the night before the second Eid, to find out whether we would be allowed or not... I think, on the government's part, that was really unfair. If it had been Christmas Eve, I don't think they would have left it so late, put it that way.”* [P23]

A key inequality that was at the forefront of these interviews was the impact of socio-economic status (SES) in which the financial impact of the pandemic was seen more clearly in these follow-up interviews for this group. Issues related to not being able to pay rent or having insecurity over work and finances was a major cause of anxiety for individuals.

*‘’ And it has sort of got to the point where I can’t pay the rent in its entirety until l get my next pension amount. And then when I take that money out of the pension, I am stuffed for the next lot because it is just a self-perpetuating shortage.”* [P13]

*‘’ I have been looking into benefits, which has been annoyingly complicated. Universal Credit is a mess, as far as I can tell, and trying to look at any benefits I might be eligible ...that hasn’t been good, at all… there is a gap there, as far as I can see, for people who suffer with mental health problems.’’* [P26]

A key gap noted by a few participants was the lack of support to those with mental health problems which were not always covered by universal credit or certain benefits which became an issue where people were having to access these benefits for the first time due to the pandemic.

Furthermore, the move to remote services and working from home further highlighted these issues of poverty and lack of access to technology or infrastructure to engage remotely. This digital move also highlighted the unequal impact on older populations who felt left behind by moving so quickly to online services as well as those who found online spaces challenging, e.g. participants with autism.

*“And then you've got internet poverty and not having the right equipment or having the right speed or having the right tech and stuff like that.”* [P17]

*‘’ Because, again, of autism, I sometimes find it hard to focus on what people say, unless there is some sort of visual connection to it. So, talking on a phone can be a bit difficult sometimes, because my brain will skip over some of the words that they’ve said. The ideal would be if everybody could just have subtitles built in. But the second best to that is being able to see somebody talking as they speak.”*

*“I worry about the elderly who don’t have access to this and you need to learn.”* [P42]

### **Physical health: Theme Summary: 7**

COVID-19 and lockdown had widespread impacts on participants’ physical health. While most participants tried to stay active at the beginning of the first lockdown, many reported leading more sedentary lifestyles and hardly going out by the time of the second interview. This was exacerbated by pre-existing physical health conditions, being hospitalised, and the winter season. As a result, many gained weight or developed bodily aches over the past months.

*“Well, I put on weight, I’ve been less active and sometimes I have neck and shoulder pain because I’m on the computer inactive.”* [P48]

Additionally, participants reported feeling isolated and lonely due to the lack of physical activity.

*‘’ I’m losing my mobility, which is absolutely freaking me out, because now the loneliness has got worse ... I can’t walk, and it’s absolutely freaking me out, because now I’m really stuck, you know.’’* [P6]

*‘I have been six months without swimming or badminton. So, that might have contributed to my stiffness”* [P13]

For others the lockdown increased stress which triggered pre-existing mental health problems and resulted in lack of sleep and physical problems.

Similar to the first interviews, participants reported they did not receive appropriate physical healthcare or felt they were not taken seriously by their GPs.

*“I would say not having the right treatment, it affects my wellbeing in terms of- I just like stick to myself. I don’t go out. I just isolate myself. I get very depressed and stuff like that.”* [P36]

Others mentioned being more reluctant to seek treatment because of their anxiety around using public transport or being treated at a hospital.

### **Perceptions of the public and policy makers: Theme Summary: 8**

Many participants mentioned disappointment in the governmental handling of the pandemic as well as the detrimental impact of social media consumption on their mental health. These issues seem to have emerged since the first round of interviews as they were barely addressed then.

Many voiced frustrations about mental health problems not being recognised and taken as seriously as physical health problems by the government.

*“I think they get side-lined by Covid to the point where they forget that people have mental health problems.”* [P46]

They hoped that the government would address the heightened detrimental impact of isolation on people with mental health problems.

*“The only thing, but it’s much more a governmental level, would be a recognising the effects of isolation during COVID-19 on people who suffer from depression. Putting in an allowance there for increased contact with a set number of people.”* [P22]

While some participants trusted the government earlier, by the time of the follow-up interview most described the guidance as illogical, inconsistent, constantly changing, as well as badly communicated. This was perceived as anxiety inducing and left many feeling isolated. As a result, some stuck to the earlier, stricter guidelines or turned to advice from experts.

*It’s the one thing that has changed in my opinion of the government since your last interview, because I’ve got angrier at them. So, I am just going with my own instincts, going with what the experts are saying”* [P27]

Additionally, participants believed that the guidance were put into place too slowly as well as were generally too liberal. Only one participant mentioned that the guidance offered reassurance to people, especially those with mental health problems.

A few participants sought information about COVID-19 on Twitter or Facebook, however, many tried to limit their exposure to social media and conspiracy theories as this was a source of anxiety, especially as people with mental health problems. One patient questioned the existent of the virus as their personal experiences diverted from the reports of high numbers of COVID deaths.

*‘There has been a lot of negative and just stupid things on Facebook that I have seen, conspiracy theories and God knows what. So, I have whittled Facebook down to about five people that I actually want to see what they are up to, you know.”* [P26]
