## Supplementary material for "What has changed in the experiences of people with mental health problems during the COVID-19 pandemic? Findings from follow-up interviews using a coproduced, participatory qualitative approach": Follow-up Interview Topic Guide

**Title of Study:** Exploring the lived experiences of loneliness and isolation with people with mental health problems during the COVID 19 pandemic in the UK

Follow-up Interview Topic Guide

Introduction

Thank you so much for agreeing to be interviewed a second time.

As you know the purpose of the interview is to find out about changes since your previous interview, to your experiences during the virus outbreak, particularly about your experiences of mental health services. Also we want to find out about changes to your experiences of feeling lonely or isolated and how they may relate to experiences of mental health problems during the virus outbreak. This will help us understand more about mental health, accessing mental health services and support, isolation and loneliness and how it relates to the virus outbreak. We will also learn more about the other challenges you have faced during the virus outbreak and what can be helpful for people experiencing mental health problems during this new situation.

There are no right or wrong answers, as everyone’s experience will be different. What you say will be kept completely confidential and we’ll anonymise the information you give us, which means that we won’t use your name or say anything that could identify you, especially in anything we write. Please be as open as you feel comfortable, and if you want to skip a question or take a break at any point, please let me know. The interview may take an hour or a bit longer, so I will ask you at several points during the interview, if you would like to take a break.

First, I’m going to ask you for some of the information about yourself that myself or my colleague asked you for before, to see if anything has changed. You don’t have to provide any information that you don’t feel comfortable sharing.

In your last interview on [date] you told me, or my colleague…

(Summary of the below information from last interview will be prepared:) You were using mental health services Y/N Has that changes since your last interview?

Types of services used: Has that changes since your last interview?

Lived with: … , in …. Has that changes since your last interview

Working, volunteering, education. Further detail they told you, ie furloughed etc Has that changes since your last interview?

·	Have you been given a new diagnosis of a mental health condition since the last interview?

·	Have you been given a new prescribed medication for mental health condition since the last interview?

•	What do you think about the current government guidance to the public regarding the virus (give examples of latest guidance at time of interview eg wearing face coverings in shops)?

·	Are you currently trying to follow the government advice (regarding the virus outbreak)?

·	If not, how long ago did you stop following the government advice regarding the virus outbreak?

Have you had any COVID-19 symptoms, or have you been diagnosed with the virus since the last interview on (date of last interview)?

1. Since your last interview, can you tell me about the main impact the virus outbreak (coronavirus /COVID-19 virus outbreak) has had on you? Currently what is the main impact on you, day to day?

Prompts: day to day impact on routine, who you saw in person/online, changes to relationships- are some stronger/weaker, practical issues, access to services and support, impact on mental health, change in symptoms over time. Any worries or concerns about getting the virus? Any impacts still being felt?

Any positive effects, e.g. new hobbies, new ways to keep in contact with people, particularly at weekends when there are less community activities available. Any impact on physical health problems, gaining weight, attending outpatient appointments etc?

2. What support from mental health services have you had in the last few months since your last interview?

Prompt: Tell us a bit more about the services you have used since your last interview, Prompts: hospital admission, Community crisis services, were any usually available services closed? Have you accessed support for your mental health from your GP?

Have you had face to face appointments? With whom, what was it like and was it any different from usual? Did they use Personal Protective Equipment (masks and gloves etc.) and did they ask you to where a mask? What was that like?

3. Were you offered any video call appointments by mental health services? Did you take up the offer? If you didn’t take up offer, why is that? If it wasn’t offered, how do you feel about it?

If Yes, took up offer of video call appointments, ask:

How were they? Were they with people you knew already or new people?

Did you experience any difficulties taking part in a video call? Was there any help you got, or think you could have got, that would make appointments by video call easier? What platform did you use? What did you think of it?

Do you think having an appointment by video call made any difference to your relationship with staff?

What would you think of having appointments by video call in future, any pros and cons?

Is there anything that would influence your willingness to have them this way?

4. Have you had any appointments by phone with mental health services?

Was it offered? If you didn’t take up offer, why is that? If it wasn’t offered, how do you feel about it?

If yes, had phone appointments, ask:

How was that? If you have had appointments by both phone and video call, how do they differ?

What is the relationship with staff like by phone? Does it make any difference to what you feel able to talk about?

What would you think of having more appointments by phone in future, any pros and cons?

5. Have you had conversations by text message, email, WhatsApp text message or any messaging tool instead of appointments?

Was it offered? If you didn’t take up offer, why is that? If it wasn’t offered, how do you feel about it?

If, yes, had conversations via text message etc, ask: How was that?

Before I move onto the next set of questions, I just want to pause to ask how you are finding the interview so far?

Would you like to take a break for about 10 minutes at this point?

6. Have you been to any online support groups?

Was it offered? If you didn’t take up offer, why is that? If it wasn’t offered, how do you feel about it?

If yes, been to online support groups, ask:

Who ran them, and what were they for? How did they compare with face to face? Do you see a role for them in future? Do you have any concerns about how it was facilitated, for example was there guidance or ground rules for people in the group?

7. Have you used any helplines?

If yes ask:

How useful was that? Do you see a role for this in future?

8. Have you used any mental health apps or websites?

If yes, ask:

What were they and how useful were they?

9. Have you had any problems getting access to support you felt you needed? Please tell us about this?

10. Since our last interview, do you think there are any ways mental health, social care or voluntary sector services could have responded better to support your mental health needs during the virus outbreak?

Prompt: what could have been done better? E.g. shift from face to face to online contact? Amount of support?

11. Are there any changes that have been made to how mental health services are delivered, that you would like to continue, or that you would not like to see continue?

Prompt: What about specific changes to the services you mentioned. What would you like to see happen? Any suggestions? What changes would you like to stay, what would you like to return to how it was before the virus outbreak. Have you been told about any long-term changes?

12. Since our last interview, what has helped support your mental health?

Prompt: friends, family, any online resources, social media, mutual support groups, mental health services, local voluntary or neighbourhood resources.

Have you either started your own projects to look out for friends and neighbours, or have you joined other initiatives, locally or larger, or online, or face to face? What impact has that had on your mental health?

13. After our last interview we gave you a list of sources of support regarding mental health, isolation and the virus outbreak. Have you used any of these resources; how did you find them if so? (as the list of resources was long, we could send it to them before hand to refer to.)

Prompts: Did you find them helpful? If yes, in what way? Were any of the resources unhelpful, or were there ones you didn’t want to try? Reasons for not wanting to try? if you made contact with them, or used their services where they helpful/unhelpful?

Before I move onto the final set of questions, I just want to pause to ask if you would like to take a break for about 10 minutes at this point?

14. Since our last interview are there changes you have made, due to the virus outbreak, in how you deal with your mental health and loneliness which you think will last beyond the end of the virus outbreak?

Prompts: mental health of loneliness self-management strategies? NHS and social care support? Support from local community groups, or peer support from other people with mental health problems? Different approaches to contacting friends and family?

15. Since our last interview, what has helped you the most to cope with the impact of the virus outbreak on your life?

Prompts: any specific resources, people, activities, any daily routine of activities?

16. Currently, do you feel lonely?

Prompts: has this changed over time during/following the virus outbreak, has your sense of belonging to the community you live in changed (neighbourhood, family and friends, communities of interest)?

17. Since our last interview, could you tell us about who you have contact with, both face-to-face, on the phone, over social media and via technology like Skype, Zoom, WhatsApp, currently? (is this different to before the virus outbreak started?)

Interviewer Prompts:

For example, are there people you see or speak to each day, every week or on a regular basis?

Do you belong to any groups or clubs? Could you tell me about your use of technology and social media?

18. Now that restrictions are changing, do you have any concerns over the medium or longer term about the impact of the virus outbreak?

Prompt: economic fall-out, dating, deaths of loved ones, restrictions on work or social life, or returning to work or socialising further shielding of loved ones above 70 or in specific health categories.

My final interview question is:

19. In the light of your experience since our last interview, what tips or advice would you give someone about coping with mental health concerns and loneliness?
